# RM-LT Trial Study Design: A Controlled Trial Exploring Remimazolam Besylate–Based Anesthesia in Enhanced Recovery After Liver Transplantation

**DOI:** 10.64898/2026.07.15.26358138

**Authors:** Xianglong Ge, He Dong, Congying Wang, Aijie Liu, Xiaojia Hao, Xiaolin Xu, Pingping Liao, Yu Wang, Boyu Kong, Lin Lyu

**Author notes:** Corresponding author: Lin Lyu.

## Abstract

**Background:** Liver transplantation is a complex surgical procedure featuring prolonged operative time and extensive surgical trauma, which results in a high anesthetic risk. Especially during anesthesia induction and the anhepatic-to-reperfusion phases, where marked hemodynamic fluctuations may readily lead to malignant cardiovascular events.

Liver transplantation is associated with numerous postoperative complications, including pulmonary complications, postoperative delirium, acute kidney injury, and delayed emergence, all of which may adversely affect patient prognosis.

Studies on remimazolam besylate (hereafter referred to as remimazolam) have suggested that it has minimal impact on patient hemodynamics. In theory, this renders remimazolam an ideal sedative agent for liver transplantation.

This study aims to verify the hypothesis that the use of remimazolam during liver transplantation can reduce the incidence of postreperfusion syndrome (PRS). In addition, we further explored the effects of remimazolam on postoperative complications in liver transplant recipients, particularly focusing on perioperative liver and kidney function, pulmonary complications, and postoperative delirium.

**Methods:** This study is a prospective randomized controlled trial. 120 participants aged 18–60 years who are scheduled to undergo liver transplantation under general anesthesia will be enrolled. In the intervention group, remimazolam besylate will be used for anesthesia induction and maintenance at doses of 0.2–0.4 mg/kg and 1–3 mg/kg/h until the end of surgery. In the control group, propofol will be used for anesthesia induction and maintenance at doses of 1–2 mg/kg and 4–12 mg/kg/h until the end of surgery.

In both groups, anesthetic drug doses or sevoflurane administration for intravenous–inhalational combined anesthesia will be adjusted based on vital signs and BIS values. All other anesthetic medications will be conducted according to the anesthetist’s preference and remain consistent.

**Discussion:** This trial will investigate whether remimazolam besylate can be used during liver transplantation to reduce the incidence of postreperfusion syndrome. It will also examine whether the drug provides potential benefits regarding perioperative complications such as postoperative delirium and acute kidney injury.

**Trial Registration:** Chinese Clinical Trial Registry,ChiCTR2500095774, registered on January 13, 2025

**Strengths and limitations of this study:** *Strengths:*

1. This study is the first clinical research on the application of this drug in liver transplantation surgeries, and it is of a prospective nature.
2. The expected results of this study can provide a safer and more advanced drug selection model for liver transplantation surgeries.

*limitations:*

1. Due to the urgency of liver transplantation surgeries, we cannot guarantee that the donor conditions are relatively consistent.
2. The general conditions of patients undergoing liver transplantation are complex and inconsistent, and we cannot predict in advance whether the baseline data of the two groups are consistent.

## Introduction

As the terminal stage of cirrhosis and various chronic liver conditions, end-stage liver disease is associated with high mortality and represents a major global public health concern, with high disability-adjusted life years (DALYs) and heavy disease burden.[1,2,3]

Liver transplantation (LT), as an effective intervention for patients with end-stage liver disease, is currently the only approach capable of providing a definitive cure and significantly prolonging survival, allowing patients to achieve favorable outcomes.[4]

### Postreperfusion Syndrome

Postreperfusion syndrome (PRS) is a severe intraoperative complication of liver transplantation, typically occurring after portal vein unclamping. It may lead to profound cardiovascular events and metabolic disturbances.[5–8].

The pathophysiological mechanisms of post-reperfusion syndrome (PRS) remain unclear, associated with metabolic acidosis, hyperkalemia, hemodynamic alterations and other relevant factors.[9–13]

Across various study centers, the reported incidence of PRS varies significantly—approximately 15–50%—due to differences in PRS definitions, treatment strategies, donor-recipient preoperative status, and surgical techniques.[13–18]

### Choice of Anesthetic Agents for Liver Transplantation

Propofol is the most common induction sedative, combined with inhalational anesthetics (e.g., sevoflurane) and opioids for maintenance and analgesia. Liver transplant patients poorly tolerate hemodynamic fluctuations, and propofol (a traditional intravenous anesthetic) is associated with a higher hypotension incidence than remimazolam; a meta-analysis by Bruno et al.[19] showed remimazolam causes less cardiopulmonary depression and fewer adverse reactions.

Anesthetic choice affects ischemia–reperfusion syndrome (PRS) incidence; propofol dose-dependently suppresses sympathetic vasoconstrictor activity, facilitating PRS.[20] Remimazolam, an ester-based benzodiazepine, induces sedation via GABA-A receptor activation. Shi et al.[21] demonstrated its protective effect against hepatic ischemia–reperfusion injury via MAPK/ERK pathway inhibition, and Nora et al.[22] confirmed its in vitro safety in human hepatocytes.

Case reports (Matsumoto et al.[23]) showed successful remimazolam-based anesthesia for allogeneic liver transplantation with stable hemodynamics, though preoperative propofol use confounded its postoperative recovery effect. The relationship between remimazolam and postoperative delirium (POD) is controversial; [24–26] recent studies[27–30] suggest no increased delirium risk, but large-scale trials are lacking.

A randomized controlled trial is urgently needed to evaluate remimazolam’s benefits in liver transplantation. This study tests whether remimazolam reduces intraoperative PRS incidence and examines its effects on perioperative organ function, pulmonary complications, and POD.

### Research Objective

The purpose of this study is to compare remimazolam and propofol through a clinical randomized controlled trial in terms of their effects on hemodynamic changes during anesthesia induction, occlusion and reopening of the superior and inferior vena cava, and reopening of the portal vein, as well as various postoperative complications, especially the incidence of postoperative delirium, ICU stay, and duration of invasive ventilation, in liver transplantation surgery. The aim is to explore the feasibility of using remimazolam besylate for general anesthesia in accelerated recovery surgery for liver transplantation.

## Methods

This protocol has been developed in accordance with the SPIRIT 2025 guidelines.

### Study Design and Objectives

This study is a prospective, single-center, single-blind, two-arm, randomized controlled trial. We plan to enroll 120 adult liver transplant recipients scheduled to undergo liver transplantation, in order to explore the application of remimazolam-based anesthesia in enhanced recovery after liver transplantation. This study will be conducted at the Affiliated Hospital of Qingdao University. The trial was registered on January 13, 2025, at https://www.chictr.org.cn/, registration number ChiCTR2500095774.

### Inclusion Criteria

1. Age 18–60 years
2. Scheduled to undergo allogeneic liver transplantation under general anesthesia
3. Provision of signed written informed consent by the participant or authorized representative

### Exclusion Criteria

1. Planned living donor liver transplantation
2. Planned split-liver transplantation
3. Planned repeat liver transplantation
4. Allergy to remimazolam
5. Severe renal insufficiency (requiring renal replacement therapy preoperatively)
6. Acute liver failure
7. Severe circulatory instability (coronary artery disease, severe arrhythmias, severe pulmonary hypertension)
8. Multiorgan transplantation
9. Preoperative acute hepatic encephalopathy or impaired consciousness

### Withdrawal Criteria

1. The participant or legal representative withdraws consent

### Criteria for Premature Discontinuation of the Study

If, during liver transplantation, the incidence of postreperfusion syndrome is significantly higher in the remimazolam group compared with the propofol group, or if postoperative delirium or other secondary outcomes adversely affecting recovery increase, the study will be terminated. If the study is terminated, enrolled participants will be informed.

The hospital ethics committee will convene every 10 cases of liver transplantation or every 6 months to review study progress and safety information. Should concerns arise regarding study performance, the committee may recommend suspension. Resumption of the study will require renewed approval from the ethics committee.

### Primary Outcome

The primary outcome is the incidence of postreperfusion syndrome, defined as a >30% decrease in mean arterial pressure compared with baseline within 5 minutes after reperfusion, lasting longer than 1 minute.

### Secondary Outcomes

1. Duration of postoperative invasive mechanical ventilation
2. ICU length of stay
3. Incidence of hypotension during anesthesia induction
4. Incidence of postoperative delirium

In addition, operative duration, blood loss, perioperative complications (type, severity, treatment details, date of occurrence, and outcome), postoperative laboratory results, and in-hospital mortality will be collected for comparison between groups.

## Trial Duration and Timeline

The trial will be conducted from February 3, 2025 to February 28, 2026 (1 year). Patient recruitment will take place during this period. Statistical analyses will be performed at the 3rd, 6th, and 9th months of recruitment, and the results will be reported to the hospital ethics committee to evaluate study safety. Statistical analysis will be completed within three month after the last patient’s data collection is finalized.

### Ethical Considerations

#### Ethics Committee Approval and Trial Registration

This study has received ethical approval from the Ethics Committee of the Affiliated Hospital of Qingdao University (Project Number: QYFYEC2024-258). The principal investigator will regularly submit study progress reports to the ethics committee. The study was registered on January 13, 2025, in the Chinese Clinical Trial Registry (Registration Number: ChiCTR2500095774).

### Written Informed Consent

For all potential study participants, the investigator must provide written informed materials explaining in detail the core objectives of the study, specific procedures, and potential benefits and risks of participation. The investigator must clearly inform every potential participant that they have the right to withdraw consent at any stage of the study. After receiving notification of organ availability, the research team will promptly provide study-related information to the participant; however, under such circumstances, potential participants may have limited time to fully consider all aspects.

Only after the participant or authorized representative signs the written informed consent form can the individual be enrolled in the study. The signed consent form will be retained as a core document of the clinical trial.

### Explanation of Organ Sources and Legality

Organs/tissue will be sourced ethically.Organs/tissue will not be sourced from executed prisoners or prisoners of conscience or other vulnerable groups. All organ donations from the deceased are non-targeted donations. To ensure fair, just, transparent and traceable organ allocation, all organs donated by the deceased will be automatically distributed through the China Organ Transplant Response System(COTRS) in accordance with national policies[31–33].The organs/tissues all come from donors who have signed an organ/tissue donation consent form in written form by themselves or all of their immediate family members.

The relevant written documents that have undergone de-identification, such as the work approval certificate issued by the research ethics committee and the donation consent form, can be obtained by contacting the corresponding corresponding author, Lin lyu, or the first author, Xianglong Ge.

### Privacy Protection

All personal information of study participants will be kept confidential. All paper and electronic materials will be securely stored by the principal investigator. All personal information related to participants will be anonymized, and the handling of sensitive information (such as biometric data) will require additional patient consent.

### Randomization and Blinding

A sealed-envelope method will be used to randomly allocate patients in a 1:1 ratio to the propofol group or the remimazolam group. Before surgery, the study coordinator will open the randomization envelope.

Because remimazolam and propofol differ in appearance, anesthesiologists and assistants will not be blinded to group assignment. They will implement the study protocol and collect intraoperative data, but they will not be informed of the study’s aims.

All patients, data collectors, outcome assessors, and statisticians will remain blinded to group allocation.

#### Unblinding

For each patient, after completion of the study (30 days postoperatively), all data will be reviewed for quality and entered into the database. Once the database is locked, unblinding (revealing group assignments) will be performed.

Postoperative follow-up will be conducted by trained data collectors who are not involved in perioperative care. Anesthesiologists and investigators will not communicate during data collection and analysis. Throughout the study, the researchers performing data analysis, medical team members, and participants will remain unaware of the treatment allocation.

## Study Interventions

### Preoperative

No premedication will be administered before the patient enters the operating room for anesthesia.

### Intraoperative Monitoring

After entering the operating room, the patient will undergo routine monitoring of pulse oxygen saturation, noninvasive blood pressure, and invasive radial arterial blood pressure. Five-lead ECG and bispectral index (BIS) monitoring will also be applied.

After anesthesia induction, the internal jugular vein will be cannulated for placement of a central venous catheter (CVC) and a sheath introducer, followed by continuous intraoperative monitoring of central venous pressure (CVP).

Nasopharyngeal temperature will be routinely monitored, and a urinary catheter will be inserted to monitor urine output. A Swan–Ganz pulmonary artery catheter may be inserted via the sheath introducer to measure pulmonary artery pressure (PAP), and transesophageal echocardiography (TEE) may be used intraoperatively when necessary.

### Anesthesia Induction

#### Propofol Group

Propofol (1–2 mg/kg), etomidate (0.2–0.6 mg/kg), sufentanil (0.1–0.3 μg/kg), cisatracurium besylate (0.15–0.2 mg/kg), or rocuronium bromide (0.6–1 mg/kg) will be administered for anesthesia induction, followed by tracheal intubation.

If a difficult airway is anticipated, succinylcholine may be used for rapid-sequence induction.

#### Remimazolam Group

Remimazolam besylate (0.2–0.4 mg/kg) will replace propofol for anesthesia induction. All other medications and induction procedures will follow the same protocol as in the propofol group.

### Anesthesia Maintenance

#### Propofol Group

A microinfusion pump will be used to continuously infuse propofol (4–12 mg/kg/h) until the end of surgery. Cisatracurium besylate (0.05–0.1 mg/kg/h) will be infused to maintain muscle relaxation. Sufentanil will be administered intermittently to ensure adequate analgesia. The anesthesiologist may adjust the concentration of sevoflurane to maintain appropriate anesthetic depth.

#### Remimazolam Group

A microinfusion pump will be used to continuously infuse remimazolam besylate (1–3 mg/kg/h) instead of propofol, and infusion will continue until the end of surgery. All other anesthetic management practices will follow the same protocol as in the propofol group.

### Intraoperative Management

#### Intraoperative ventilation management

1. Mechanical Ventilation Strategy
2. Tidal volume (Vt): 6–8 mL/kg
3. PEEP: 3–5 cmH₂O
4. Plateau pressure: <30 cmH₂O
5. Respiratory rate (f): 10–16 breaths per minute
6. FiO₂: 50%–70%

#### General Intraoperative Management

Anesthetic depth will be adjusted to maintain BIS values between 45–60. Albumin and crystalloid fluids will be routinely infused. Arterial blood gas analysis will be performed at least once every hour. Based on individual patient needs, third-generation cephalosporins or carbapenems will be administered intravenously for infection prophylaxis.

Basiliximab (Simulect) or rabbit anti-human thymocyte immunoglobulin (ATG), combined with corticosteroids, will be administered intraoperatively to suppress immune responses. Methylprednisolone sodium succinate will be administered intravenously before graft reperfusion.

Transfusion of blood products will be guided by anesthesiologists’ clinical experience and judgment, maintaining hemoglobin levels at ≥7 g/L. Plasma, platelets, cryoprecipitate, fibrinogen, and prothrombin complex concentrate will be used to manage coagulopathy.

Vasopressors and inotropes will be administered when necessary to maintain relative hemodynamic stability.

#### Anesthesia Emergence and Postoperative Management

At the end of surgery, infusion of propofol/remimazolam and neuromuscular blockers will be discontinued. Analgesics (e.g., sufentanil) will be administered as needed. Before leaving the operating room, the concentration of sevoflurane will be reduced or discontinued based on vital signs.

It is routine at our center to transfer post–liver transplantation patients to the liver transplant ICU for postoperative care. Patients will receive level-one monitoring upon ICU admission, with close observation of vital signs. No special Concomitant care after the operation.

Sedation and analgesia will be assessed hourly, and interventions will be applied based on postoperative status. Sedative medications such as propofol, midazolam, and dexmedetomidine may be used according to individual needs, and butorphanol tartrate will be administered for postoperative analgesia.

Our center does not recommend patient-controlled analgesia to avoid potential adverse reactions.

#### Care of organ donors

Before organ donation, a strict assessment ensures legality, voluntariness, and standardization in line with national regulations. Before death is declared, patient care remains the priority. An OPO coordinator assesses donation potential while the ICU provides basic life support. OPO coordinators empathetically engage the family, explain donation, respect their beliefs, and obtain written consent. After family agreement, the team plans end-of-life care, death determination, and organ retrieval with humanity and dignity. The patient is moved from the ICU to the operating room for life support removal and organ recovery. The OPO coordinator places catheters for blood pressure monitoring, organ perfusion, medication, and internal environment stability. Life support removal is patient-centered, with pain relief and sedation. After removal, death is confirmed by monitoring and physical exam, followed by a 2–5 minute observation period without resuscitation. The OPO then handles the body: suturing, tube removal, cleaning, and appearance restoration. Family may stay or wait in a dedicated area, receiving psychological counseling. The kindness of the donor and their family should be formally thanked and cared for.

## Data Collection

The study design is shown in the flowchart in Fig.1, and the study schedule is presented in Fig.2.

### Preoperative Data Collection

After the participant agrees to join this study and signs the paper-based informed consent form, baseline data will be collected:

1. Demographic characteristics (age, sex)
2. Past medical history
3. Admission diagnosis (reason for undergoing liver transplantation)
4. Physiological indicators (coagulation-related laboratory tests, hemoglobin, myocardial injury markers, serum creatinine, urea, direct/indirect bilirubin, etc.), MELD score, Child–Pugh score
5. Smoking and drinking history, drug/food allergy history
6. Donor liver characteristics, including cause of death, height, weight, graft quality, ischemia time, and liver disease history

### Intraoperative Data Collection

1. Date of surgery, surgical procedure, ASA classification, duration of surgery and anesthesia
2. Invasive arterial pressure, oxygen saturation, and heart rate upon entry into the operating room
3. Types and dosages of intraoperative anesthetic medications
4. The types and dosages of drugs used for anesthesia induction, as well as the hemodynamic parameters within 10 minutes after anesthesia induction
5. Invasive arterial pressure, pulmonary artery pressure, and heart rate before portal vein unclamping, at the moment of unclamping, and at 10 s, 20 s, 30 s, 40 s, 50 s, 60 s, 2 min, 3 min, and 5 min after unclamping; total doses of vasopressors administered within the first 5 minutes via bolus or infusion
6. Intraoperative fluid management parameters, including crystalloid/colloid intake, blood loss, ascites volume, urine output, types and volumes of blood products infused
7. Vital signs from post-induction until the end of surgery, including BIS, noninvasive/invasive arterial pressure, heart rate, oxygen saturation, pulmonary artery pressure, central venous pressure, and hemodynamic parameters obtained using the Flotrac sensor (CO, SV, SVV, SVRI)
8. liver status, including cold ischemia time, laboratory tests, etc

### Postoperative Data Collection

After surgery, patients will routinely be transferred to the liver transplant ICU for monitoring. Postoperative follow-up and data collection will begin 2 hours after surgery. ICU bedside assessments will be conducted at postoperative hours 2, 6, 12, 24, and 48. Laboratory tests and ICU hospitalization data will be collected from electronic medical records or through telephone follow-up by data collectors.

Collected postoperative data include:

1. RASS score and delirium assessment (using the CAM-ICU method)
2. Types and dosages of sedatives and analgesics
3. Duration of ICU and hospital stay, and duration of invasive mechanical ventilation
4. Laboratory test results within 7 days postoperatively (creatinine, bilirubin, ALT, AST, BNP, high-sensitivity troponin)
5. Incidence of postoperative complications during hospitalization
6. All-cause mortality within 30 days after surgery

### Adverse Events

In this study, the medications investigated are commonly used in routine anesthetic practice at similar dosages, and no drug-related adverse events have been reported in the preliminary experiment. However, adverse events may still occur during routine anesthesia and affect participant health.

If any adverse event related to the study medications occurs, anesthesiologists will provide emergency management according to standard practice and report the event to the investigators for documentation.

All adverse events (AEs) and serious adverse events (SAEs) will be managed, recorded, and followed up to ensure that the trial does not pose health risks to participants.

## Statistics and Data Management

### Statistical Analysis

Each participant underwent an Intention-to-Treat Analysis.Statistical analysis will include analyses of numerical variables, categorical variables, and time-to-event variables. Numerical variables, including postoperative laboratory results, will be expressed as mean (standard deviation) or median (minimum, maximum; or interquartile range). The missing data will not be included in the data analysis, and will be specially marked in the database and the analysis.

Categorical variables will be presented as numbers and percentages, and comparisons between groups will be performed using the χ² test.

The Shapiro–Wilk test will be used to determine whether continuous variables follow a normal distribution. Normally distributed variables will be expressed as mean ± standard deviation, and differences between groups will be compared using the t test. Non-normally distributed variables will be expressed as median (interquartile range), and differences will be analyzed using the Wilcoxon rank-sum test. Furthermore, based on the different characteristics of the patients, subgroup analyses will be conducted according to factors such as age, gender, and cause of the disease.

Vital signs measured at multiple time points will be analyzed using repeated-measures ANOVA, with Bonferroni correction applied for post-hoc testing. All statistical analyses will be conducted using SPSS Statistics version 29.0 (IBM Corp.) and R Studio 4.2.2 (RStudio). A two-sided P < 0.05 will be considered statistically significant.

### Sample Size Calculation

Based on data from our preliminary experiment and published literature, the incidence of PRS during anesthesia with propofol is approximately 43%, while the incidence during anesthesia with remimazolam is approximately 15%.

Assuming α = 0.05 and power = 0.90, the calculated sample size is 50 cases per group. Considering a 20% attrition rate, the final sample size will be 60 participants per group.

### Data Management

Original data include metadata and data recorded on the Case Record Form (CRF). Metadata will be transcribed into CRFs. Online quality control and real-time quality control will be conducted using an internet-based Electronic Data Capture (EDC) system.

Paper-based CRFs will be used to record all study data. Investigators will ensure that CRFs are completed accurately, promptly, and thoroughly. Supervisors/study coordinators will oversee study conduct. Completed CRFs will be entered and managed by personnel responsible for data statistics.

After each participant completes the study (30 days postoperatively), all data will be reviewed to ensure quality and entered into the database. The database will then be locked, and unblinding will be performed.

All research data involving participant confidentiality will be securely stored, and the EDC system will retain data for 10 years after the trial ends. Original signed informed consent forms will be stored at each study site.

For participants who interrupted or deviated from the intervention plan, we will only collect the reasons for their withdrawal from the study. All other data will be deleted.

The data monitoring committee is composed of the initiators of this study and the designated data managers. It does not intervene in any aspect of the research. The data manager will conduct regular analyses of data availability and identify any issues that may affect the final data analysis. They will promptly report these findings to the research initiator.

### Data Availability Statement

The raw data of this study will be publicly released on May 27, 2026, and will be available in ResMan (https://www.medresman.org). The formal research report will be submitted to the trial registry and the relevant findings will be published in a journal in the form of a paper.

### Quality Control

Before the study begins, all participating personnel will undergo standardized training. Training will be conducted by the principal investigator and will include:

1. Study protocol
2. Related standard operating procedures (SOPs)
3. Guidance on CRF completion and data interpretation
4. Notes regarding study conduct (e.g., documentation and reporting of adverse events and complications)

If revisions to the study protocol are required based on study progress, training will be updated accordingly to ensure synchronized understanding.Any significant modifications to the trial protocol will be promptly communicated to the research registration authority, the ethics committee, and all the researchers and participants involved in this study.

All study personnel must strictly follow the study protocol throughout the trial. All expected and unexpected findings shall be accurately documented to ensure the reliability of study conclusions.

Statistical analysts will conduct ongoing data quality control throughout the study. If data quality issues arise, they will promptly report them to the principal investigator and provide corrective measures.

## Discussion

The RM-LT trial is a prospective, single-center, single-blind, two-arm, randomized controlled trial enrolling a total of 120 patients aged 18–60 years undergoing liver transplantation. This study will provide baseline characteristics of this surgical population and evaluate whether remimazolam, as a benzodiazepine with relatively weak hemodynamic effects, can effectively reduce the incidence of postreperfusion syndrome during liver transplantation.

Additionally, it will explore whether remimazolam may contribute to enhanced recovery after liver transplantation by reducing postoperative delirium, lowering the incidence of perioperative complications, and shortening ICU length of stay.

The results of this study will provide a new paradigm for anesthetic drug selection in adult liver transplantation and promote the application of more appropriate sedative agents for liver transplant anesthesia.

As one of the largest liver transplantation centers in China, our center completes a nationally leading number of technically advanced liver transplant surgeries, providing a large number of potential research participants. We also possess well-established surgical, anesthetic, and nursing teams, which will reduce heterogeneity in the medical care participants receive.

Furthermore, the rigorous design of this study allows us to draw more accurate conclusions and clarify whether remimazolam can benefit patients during the perioperative period of liver transplantation.

Although the study is rigorous and the procedures are clearly defined, the timing of donor liver procurement is unpredictable, and liver transplantation is often urgent. Therefore, assigning a designated anesthesiologist to every participant in advance is impractical. To minimize differences caused by individual practice habits among anesthesiologists, we will assign two anesthesiologists to each enrolled participant, with anesthesia randomly performed by one of the two.

Additionally, this study is conducted at a single center, which may limit the generalizability of the findings. At our center, the primary indications for liver transplantation include decompensated hepatitis B cirrhosis, alcoholic liver disease, and hepatocellular carcinoma, which may differ significantly from the surgical indications in other regions.

The advantages of this study include:

1. As a prospective, single-center, single-blind, two-arm, randomized controlled trial, it will be the first to provide clinical evidence regarding the potential benefits of remimazolam in liver transplantation anesthesia.
2. The study will collect comprehensive perioperative data across all phases, allowing for rigorous and reliable conclusions.
3. The raw data will be publicly released after the study, providing a valuable data source for this type of surgery and promoting scientific research in related fields.

### Trial Status

This trial was registered on January 13, 2025, in the Chinese Clinical Trial Registry (Registration Number: ChiCTR2500095774). This research has completed the participant recruitment stage. The data analysis phase will commence on May 15th.

Protocol version: 1.0.

## Supporting information

SPIRIT 2025 guidelines

## ethics approval statement

This study has received ethical approval from the Ethics Committee of the Affiliated Hospital of Qingdao University (Project Number: QYFYEC2024-258).

## patient consent statement

A copy of the informed consent obtained from the patients is included as Appendix 1.

## permission to reproduce material from other sources

The authors confirm that no copyrighted materials from other publications have been reproduced in this work.

## clinical trial registration

The study was registered on January 13, 2025, in the Chinese Clinical Trial Registry (Registration Number: ChiCTR2500095774).

## Public involvement

This study did not involve any patients or the public in the details or plans of the trial design, implementation, or reporting process.

---

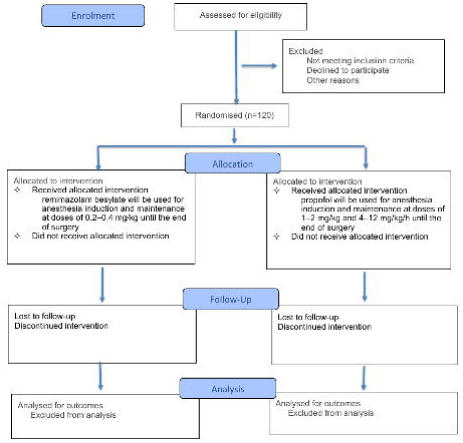

