## Supplementary material for "RM-LT Trial Study Design: A Controlled Trial Exploring Remimazolam Besylate–Based Anesthesia in Enhanced Recovery After Liver Transplantation": fig.2

| | screening period | baseline | postoperative hospitalization | | | | | | | |
|---|---|---|---|---|---|---|---|---|---|---|
| | | surgery | day1 | day2 | day3 | day4 | day5 | day6 | day7 | discharge |
| **inclusion and exclusion criteria** | × | | | | | | | | | |
| **informed consent** | × | | | | | | | | | |
| **basic information** | × | | | | | | | | | |
| **Preoperative laboratory tests** | × | | | | | | | | | |
| **donor situation** | × | | | | | | | | | |
| **Information related to surgery and anesthesia** | | × | | | | | | | | |
| **laboratory test** | × | | × | × | × | × | × | × | × | |
| **postoperative systemic complications** | | | × | × | × | × | × | × | × | |
| **ICU and overall hospitalization statistics** | | | | | | | | | | × |
